# Artificial Intelligence Applications for COVID-19 in Intensive Care and Emergency Settings: A Systematic Review

**DOI:** 10.1101/2021.02.15.21251727

**Authors:** Marcel Lucas Chee, Marcus Eng Hock Ong, Fahad Javaid Siddiqui, Zhongheng Zhang, Shir Lynn Lim, Andrew Fu Wah Ho, Nan Liu

## Abstract

**Background:** Little is known about the role of artificial intelligence (AI) as a decisive technology in the clinical management of COVID-19 patients. We aimed to systematically review and critically appraise the current evidence on AI applications for COVID-19 in intensive care and emergency settings, focusing on methods, reporting standards, and clinical utility.

**Methods:** We systematically searched PubMed, Embase, Scopus, CINAHL, IEEE Xplore, and ACM Digital Library databases from inception to 1 October 2020, without language restrictions. We included peer-reviewed original studies that applied AI for COVID-19 patients, healthcare workers, or health systems in intensive care, emergency or prehospital settings. We assessed predictive modelling studies using PROBAST (prediction model risk of bias assessment tool) and a modified TRIPOD (transparent reporting of a multivariable prediction model for individual prognosis or diagnosis) statement for AI. We critically appraised the methodology and key findings of all other studies.

**Results:** Of fourteen eligible studies, eleven developed prognostic or diagnostic AI predictive models, all of which were assessed to be at high risk of bias. Common pitfalls included inadequate sample sizes, poor handling of missing data, failure to account for censored participants, and weak validation of models. Studies had low adherence to reporting guidelines, with particularly poor reporting on model calibration and blinding of outcome and predictor assessment. Of the remaining three studies, two evaluated the prognostic utility of deep learning-based lung segmentation software and one studied an AI-based system for resource optimisation in the ICU. These studies had similar issues in methodology, validation, and reporting.

**Conclusions:** Current AI applications for COVID-19 are not ready for deployment in acute care settings, given their limited scope and poor quality. Our findings underscore the need for improvements to facilitate safe and effective clinical adoption of AI applications, for and beyond the COVID-19 pandemic.

## 1 Introduction

The ongoing coronavirus disease 2019 (COVID-19) pandemic has challenged healthcare systems and healthcare practitioners worldwide. Intensive care units (ICU) and emergency departments (ED) in badly afflicted areas have been overwhelmed by the surge in patients suspected or diagnosed with COVID-19 ^1-3^. This exerts significant pressure on healthcare resources, necessitating novel diagnostics and care pathways to rationally deploy scarce emergency and intensive care healthcare resources. Current strategies and recommendations on clinical management and resource rationalisation draw on past pandemic experiences and expert recommendations ^3-5^; however, there has been growing interest in novel applications of artificial intelligence (AI) to assist in the COVID-19 response within these settings.

AI is commonly defined as the use of computational methods to mimic human intelligence. Machine learning and deep learning are branches of AI which focus on automatic improvement of computer programmes through experience ^6,7^. Regression models, such as logistic, linear, or Cox regression, are simple forms of machine learning which already have longstanding use in medical research. More advanced machine learning, including random forest models, neural networks, or support vector machines, are also becoming more common in the medical literature, introducing more complex and diverse applications of AI. In intensive care and emergency settings, AI applications have assisted with automated patient monitoring ^8-11^, prognostication ^12^, and optimisation of staffing allocations ^13-16^. Given the unprecedented volume of COVID-19 patients, recent reviews have also identified resource optimisation of ICU beds as a potentially significant application of AI ^17,18^.

Earlier systematic reviews have identified significant issues in the quality and reporting of predictive models for COVID-19 diagnosis and prognosis ^19^ and AI applications for classifying COVID-19 medical images ^20^. Shillian et al.’s ^21^ systematic review of machine learning studies in pre-COVID-19 ICUs reported similar issues, such as limited sample size and poor validation of predictions. However, no study has evaluated the scope and quality of all available AI applications in intensive care and emergency settings. This gap in knowledge precludes valuable improvements to the development and deployment of AI applications in these settings. We aimed to systematically review and critically appraise the current evidence on AI applications for COVID-19 in intensive care and emergency settings, focussing on methods, reporting standards, and clinical utility.

## 2 Methods

We reported this systematic review according to the Preferred Reporting Items for Systematic Reviews (PRISMA) guidelines (Additional file 1). A review protocol was developed but was not publicly registered.

### 2.1 Search strategy and selection criteria

We searched six databases, PubMed, Embase, Scopus, CINAHL, IEEE Xplore, and ACM Digital Library, by combining search terms related to AI, COVID-19, and intensive care or emergency settings. For brevity, the search strategy showing only the first three terms in each concept set is as follows: ((“Artificial intelligence” OR “Deep learning” OR “Machine learning” OR …) AND (“COVID-19” OR “Coronavirus disease 2019” OR “2019-nCoV” OR …) AND (Emergency OR “ED” OR “intensive care” OR …)). The complete search strategy can be found in Additional file 2. We also screened the reference lists of included articles to identify additional relevant studies. We included articles that met the following criteria: (1) applied AI; (2) investigated COVID-19 operations of ICU, ED, or emergency medical services (EMS) or analysed data from COVID-19 patients in the ED or within a prehospital setting, COVID-19 patients requiring intensive care (admission to the ICU, mechanical ventilation, or a composite including either of these outcomes), or the healthcare workers treating these patients, including ED or ICU physicians and nurses as well as paramedics; and (3) were original, peer-reviewed research articles. For this review, artificial intelligence only encompassed conventional machine learning algorithms such as random forest models, neural networks, or support vector machines. Multivariable logistic regression predictive models (including ridge and least absolute shrinkage and selection operator (LASSO) regression) were excluded. No restrictions were placed on the language of articles.

### 2.2 Literature selection and data extraction

We conducted an initial search on 30 August 2020 and updated the search results on 1 October 2020. Articles were screened by title, abstract, and, if ambiguous, full text by two independent reviewers (MLC and NL). Subsequently, the two reviewers (MLC and NL) independently extracted data using a standardised data extraction form. Discrepancies in article selection and data extraction were resolved between reviewers through discussion.

We extracted the following data for all included articles: country of study population, outcome predicted, sample size of the training and validation datasets, AI algorithms used, discrimination (e.g. C-index, accuracy) and calibration (e.g. calibration slope, Brier loss score) of models on the strictest form of validation, features included in the final model, and transparent reporting of a multivariable prediction model for individual prognosis or diagnosis (TRIPOD) study type ^22^, if applicable.

### 2.3 Data analysis

For studies including multivariate AI predictive models, we evaluated the risk of bias within the study methodology using prediction model risk of bias assessment tool (PROBAST) ^23^. PROBAST is a structured tool comprising 20 signalling questions for assessing the risk of bias and applicability across the four domains of participants, predictors, outcome, and analysis. Applicability of included studies was not assessed as our study was not concerned with a specific application of AI predictive models. In lieu of specific reporting standards for AI studies at the time of study conception ^24^, we assessed the reporting quality of multivariable predictive modelling studies using an adaptation of Wang et al.’s ^25^ modified TRIPOD statement ^26^ for AI models (Additional file 3). For all other studies, we summarised the study methodology, including data sources, application of AI, and validation methods, as well as the key findings of the study.

## 3 Results

### 3.1 Study characteristics

From our search of the six databases, 14 studies were included in this review (Figure 1). Table 1 presents the main characteristics of the study. 11 of the 14 studies investigated predictive models and were assessed according to PROBAST and TRIPOD: eight studies developed prognostic models ^27-34^ and three studies developed diagnostic models ^35-37^. Of the remaining three studies, two evaluated the prognostic potential of existing AI-based lung segmentation software (without integration into a multivariate predictive model) ^38,39^ and one investigated an AI-based system for resource optimisation in the ICU ^40^. Eleven studies used patient data collected from the ICU and four studies used data from the ED. No study collected data from the prehospital setting, despite including prehospital-related search terms in the search strategy.

**Table 1.**
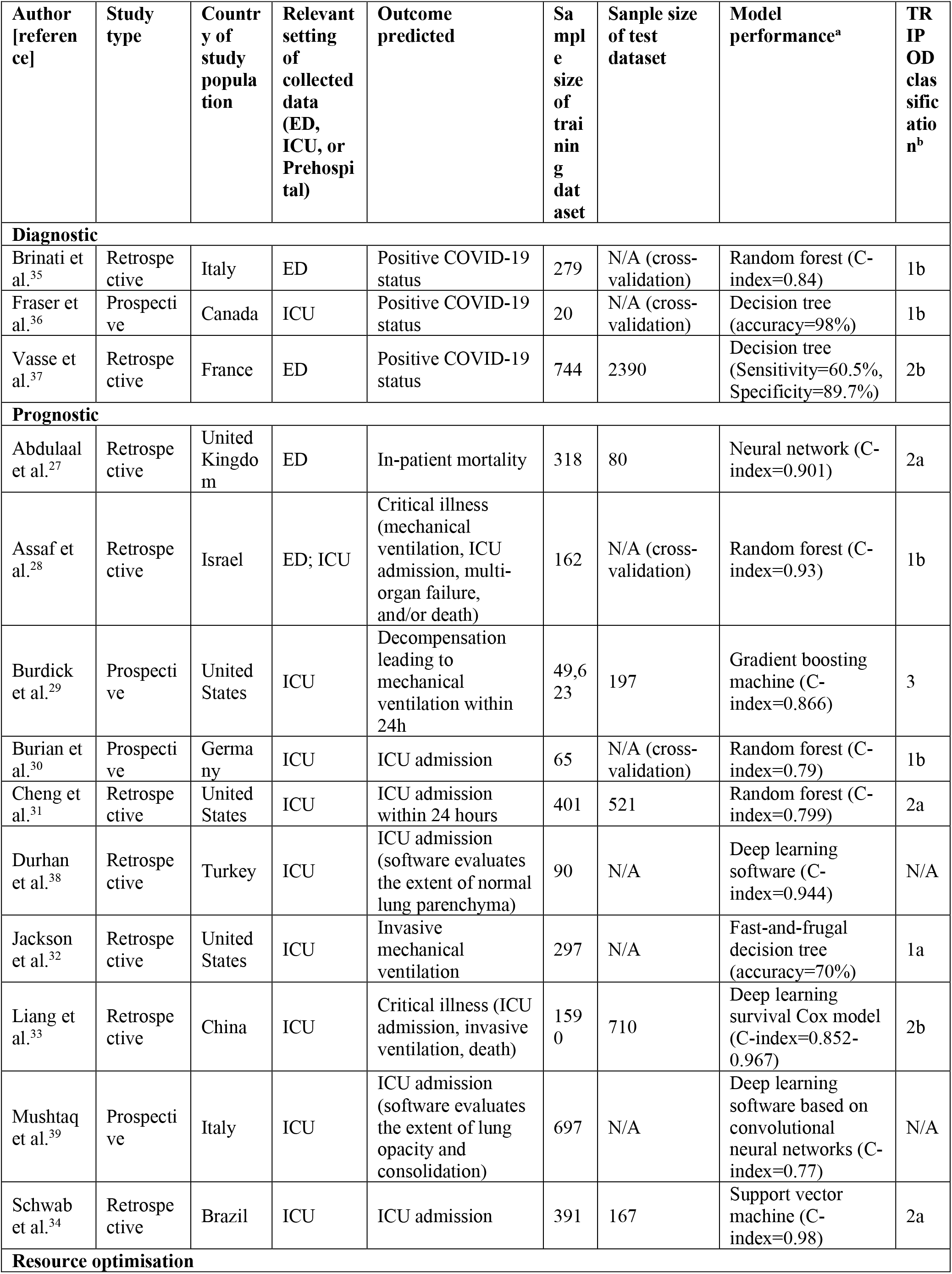
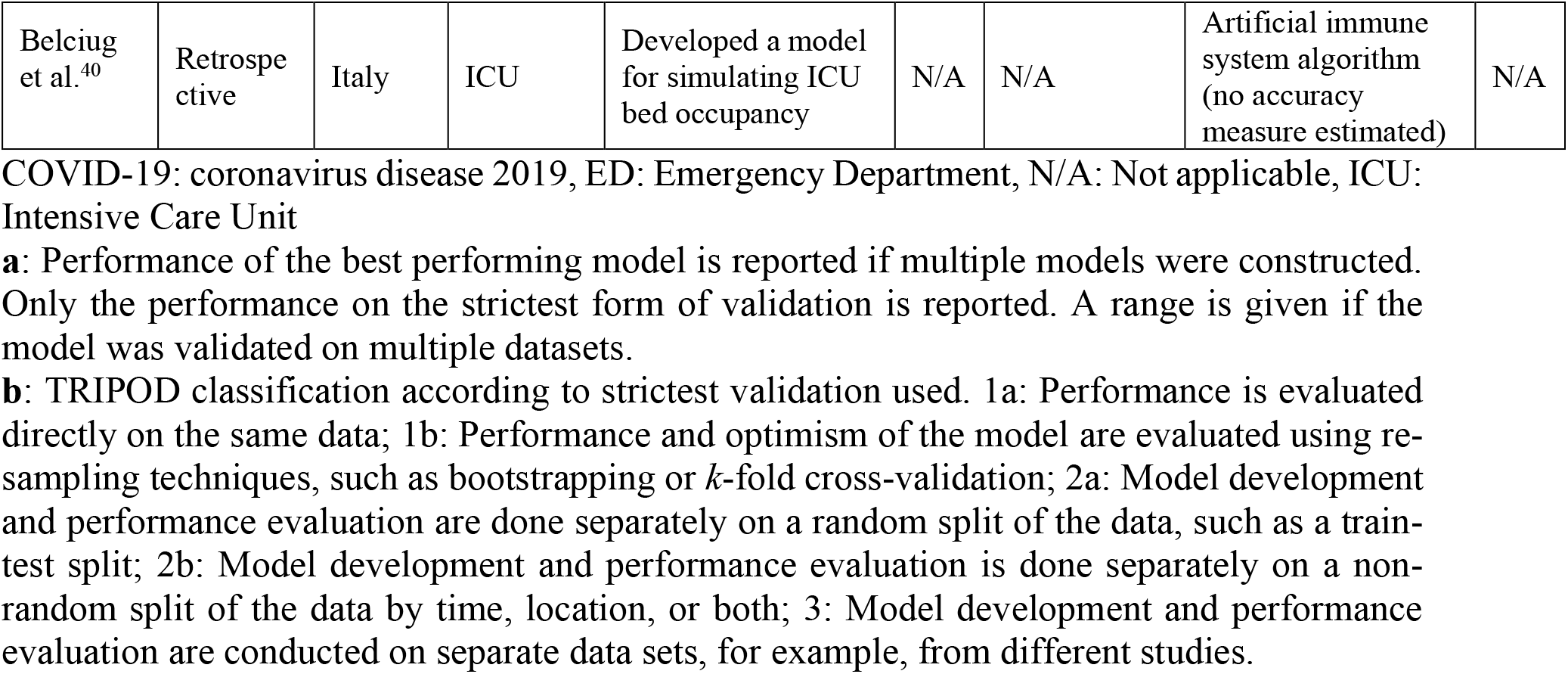
Main study characteristics.

**Figure 1:**
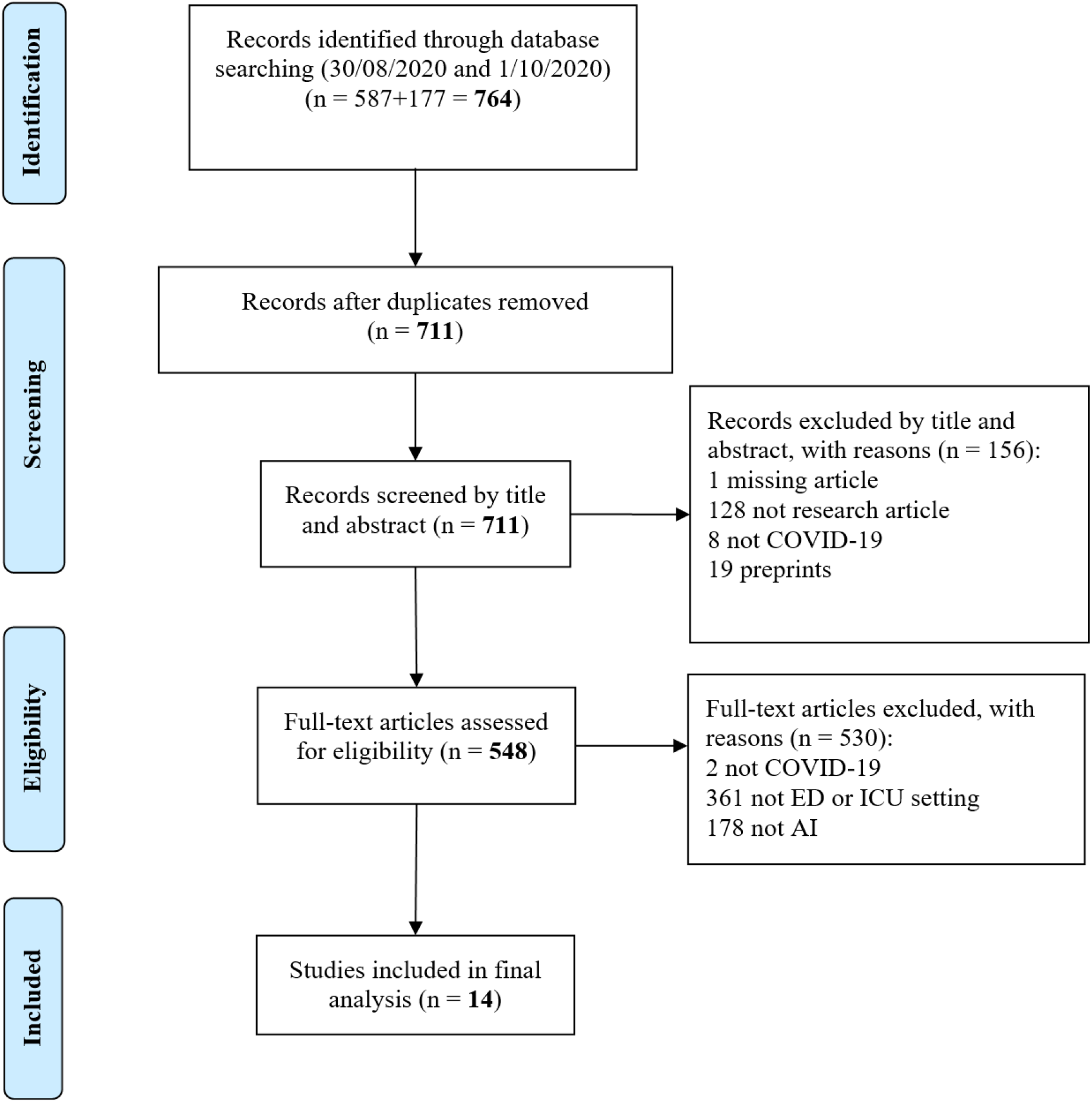
PRISMA flow diagram.

In terms of country of study, Italy (*n*=3) and United States (*n*=3) were represented by more than one study, while Brazil, Canada, China, France, Germany, Israel, Turkey, and the United Kingdom had one study each.

According to the TRIPOD classification of predictive models, two studies were classified as Type 2b (validation using a non-random split of data by time and/or location), three studies as Type 2a (validation using a random split of data such as a train-test split), four studies as Type 1b (validation using re-sampling techniques such as bootstrapping or k-fold cross-validation), and one study as Type 1a (no validation, only evaluation of apparent model performance on the same training dataset). One study that conducted development and validation using data from separate studies was considered Type 3.

### 3.2 Risk of bias

Table 2 presents the risk of bias assessment of AI predictive models according to PROBAST. All 11 predictive modelling studies had a high overall risk of bias. Two out of 11 studies had an unclear risk of bias within the participant domain. Unclear risk of bias in the participant domain was mainly due to ambiguous exclusion criteria that may lead to the study population not being representative of the intended target population ^33,34^.

**Table 2.** PROBAST (prediction model risk of bias assessment tool) assessment of predictive modelling studies.

| Author | Risk of bias according to PROBAST domain |  |  |  |  |
| --- | --- | --- | --- | --- | --- |
|  | Participants | Predictors | Outcomes | Analysis | Overall |
| **Diagnostic** |  |  |  |  |  |
| Brinati et al.<sup>35</sup> | Low | Low | Low | High | High |
| Fraser et al.<sup>36</sup> | Low | Unclear | Low | High | High |
| Vasse et al.<sup>37</sup> | Low | Low | Low | High | High |
| **Prognostic** |  |  |  |  |  |
| Abdulaal et al.<sup>27</sup> | Low | Low | Low | High | High |
| Assaf et al.<sup>28</sup> | Low | Low | Unclear | High | High |
| Burdick et al.<sup>29</sup> | Low | Low | Unclear | High | High |
| Burian et al.<sup>30</sup> | Low | Low | Low | High | High |
| Cheng et al.<sup>31</sup> | Low | High | Unclear | High | High |
| Jackson et al.<sup>32</sup> | Low | High | High | High | High |
| Liang et al.<sup>33</sup> | Unclear | High | High | High | High |
| Schwab et al.<sup>34</sup> | Unclear | Unclear | Unclear | High | High |
| Vasse et al.<sup>37</sup> | Low | Low | Low | High | High |

All three studies at a high risk of bias in the predictor domain were prognostic. Two studies ^32,33^ used retrospective, multicentre data and were at risk of bias from varying methods of predictor assessment at different centres. The remaining study ^31^ obtained predictor data from the most recent assessments available, instead of assessing predictors at the intended time of use. Two studies did not report adequately on the assessment of computed tomography (CT) ^36^ or other features ^34^, resulting in an unclear risk of bias.

Two and four out of 11 studies were at high and unclear risk of bias within the outcome domain, respectively. In many prognostic studies ^28,29,31,34^, the criteria for ICU admission and blinding of outcome determination to predictor variables were often not reported, leading to an unclear risk of bias.

Within the analysis domain, all eleven studies had insufficient outcome events per variable (EPV) (<20 EPV for model development studies and <100 for model validation studies) leading to a high risk of bias. Furthermore, no study reported on model calibration and only two studies ^34,35^ appropriately handled and reported on missing data. Prognostic predictive models were particularly at risk of inadequately accounting for, or reporting on, censored patients who were still hospitalised without the outcome (e.g. ICU admission) at the end of the study period. Only one study appropriately accounted for censored data by combining deep learning techniques with traditional Cox regression ^33^.

### 3.3 Adherence to reporting standards

The modified TRIPOD checklist comprised 25 terms, including 17 terms for reporting of methods and eight terms for results. Figure 2 describes the adherence of studies to reporting standards, as assessed by the modified TRIPOD checklist. Studies reported on a median of 48% (IQR: 48-59%) of relevant TRIPOD items, with 10 of 25 TRIPOD items having 50% adherence or less. Additionally, the following eight TRIPOD items had 25% adherence or less: reporting on treatments administered to study participants (item 5c), blinding of outcome and predictor assessment (items 6b and 7b), study size determination (item 8), reporting on characteristics of study participants, including proportions of participants with missing data (item 13b), reporting of unadjusted associations between predictors and outcomes in multivariable logistic regression models (item 14b), explanation of how to use the prediction model (item 15b), and calibration and method of calibration (adjusted item 16b).

**Figure 2:**
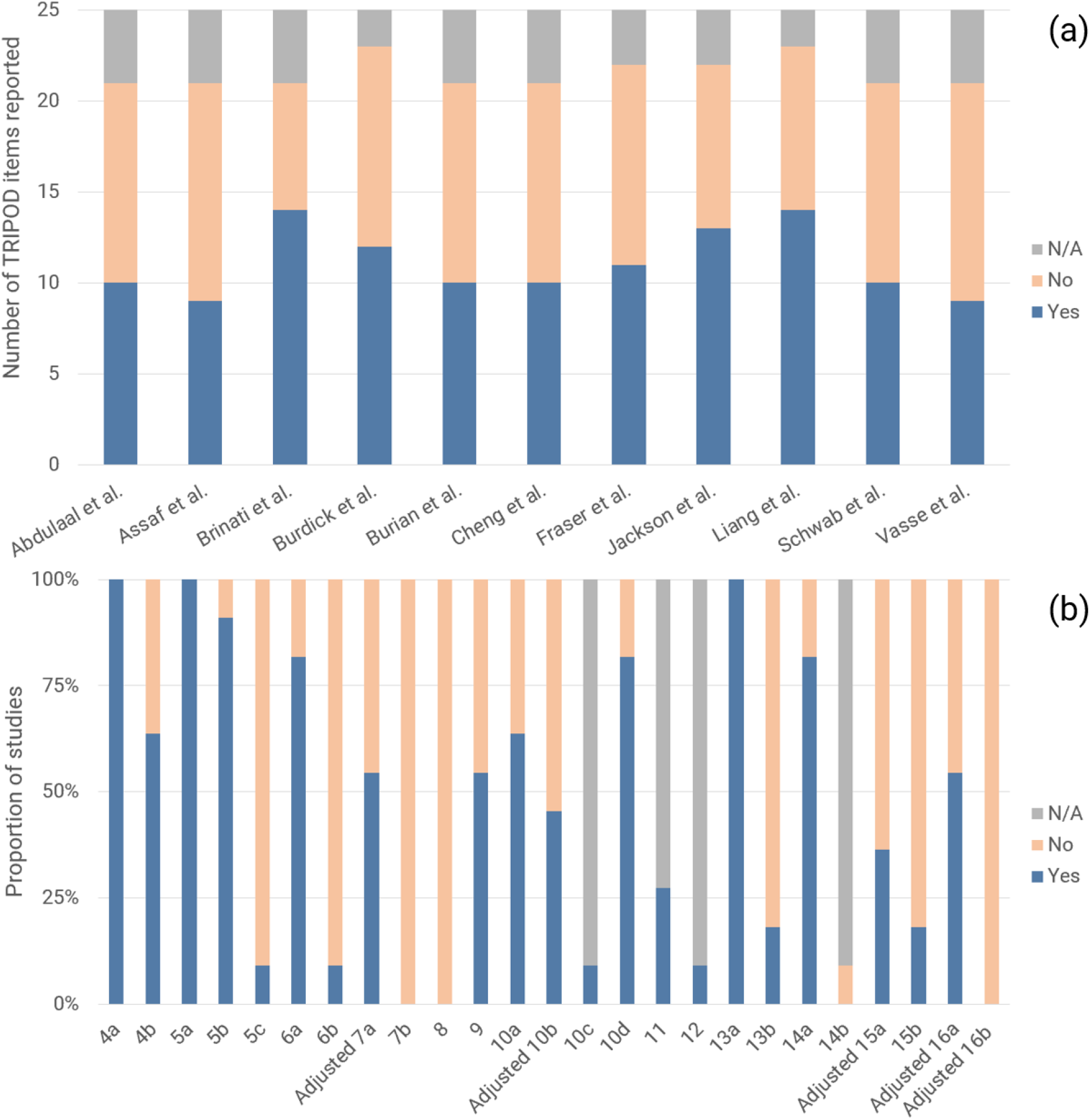
(a) Number of TRIPOD items reported per study and (b) Proportion of studies reporting on each TRIPOD item.

### 3.4 Diagnosis

Three studies investigated diagnostic AI predictive models; two studies developed models to predict the outcome of COVID-19 status at admission to the ED. Only one study was externally validated: Vasse et al. ^37^ developed a decision tree based on cellular population data using Random Forest for feature selection (accuracy=60.5%). Brinati et al.’s ^35^ Random Forest model (C-index=0.84, accuracy=82%) and Three-Way Random Forest model (accuracy=86%) achieved better performance but was validated using weaker *k*-fold cross-validation. Both studies included leucocyte or a leucocyte sub-population count as a predictor in their final model.

The third study ^36^ developed a decision tree for determining COVID-19 infection status in the ICU based on plasma inflammatory analyte features selected by a random forest classifier. On five-fold cross-validation, this classifier achieved an accuracy of 98%.

### 3.5 Prognosis

Most studies on prognostic AI predictive models (9/10, 90%) predicted ICU admission, mechanical ventilation, or a similar composite outcome of severe or critical illness. Collectively, such studies reported C-indices between 0.79-0.98. Liang et al’s ^33^ Deep Learning Survival Cox model had the largest training cohort of 1590 patients and achieved a C-index of 0.890, 0.852, and 0.967 when externally validated on cohorts of 801, 305, and 73 patients from Wuhan, Hubei, and Guangzhou, respectively. Schwab et al.’s ^34^ support vector machine achieved a superior C-index of 0.98 on a weaker internal validation and a smaller sample size for testing model performance.

The artificial neural network trained by Abdulaal et al. ^27^ using data collected at ED admission (C-index=0.901) was the only prognostic AI model developed to predict in-hospital mortality in COVID-19 patients.

Apart from predictive modelling, Durhan et al. ^38^ and Mushtaq et al. ^39^ evaluated the prognostic utility of two separate deep learning-based software that determine the normal lung proportion and total lung involvement, respectively. Scores obtained from each software achieved a C-index of 0.944 and 0.77 for predicting ICU admission, respectively. While multivariate predictive models were not developed, both studies were subject to similar issues in development and reporting, including ambiguous criteria for ICU admission, inappropriate handling of missing data using complete-case analysis, and lack of reporting on treatments received by participants and on blinding of the outcome.

### 3.6 Other applications

Apart from diagnostic and prognostic applications, Belciug et al. ^40^ utilised an Artificial Immune System algorithm, a type of evolutionary AI algorithm, to optimise a queueing model for simulating hospital bed allocation in the ICU. The final model, intended as a tool for hospital managers, proposes an optimal admission rate and number of beds while balancing the costs associated with increasing capacity and refusing patients. The model was applied to ICU data published by the Ministry of Health of Italy and estimated a minimum rejection rate of 3.4% and 1.7% of patients requiring ICU admission from 13 March 2020 to 23 March 2020 (average daily volume of 200 patients) and 23 March 2020 to 30 March 2020 (average daily volume of 63 patients), respectively. However, these estimates were not validated.

## 4 Discussion

Our study is the first systematic review of AI applications for COVID-19 in intensive care and emergency settings. Applications were largely limited to diagnostic and prognostic predictive modelling, with only one study investigating a separate application of simulating ICU bed occupancy for resource optimisation. Due to high risk of bias, inadequate validation, or poor adherence to reporting standards in all reviewed studies, we have found no AI application for COVID-19 ready for clinical deployment in intensive care or emergency settings.

Among the reviewed articles, we found a limited range of AI applications being studied within intensive care and emergency settings. An exploratory review identified early detection and diagnosis, resource management of hospital beds or healthcare workers, and automatic monitoring and prognostication as possible applications of AI for the COVID-19 pandemic ^17^. However, current applications within the reviewed articles mainly comprised prognostic models for critical illness or diagnostic models to predict COVID-19 status, none of which are ready for clinical use. Only one preliminary study by Belciug et al. ^40^, which lacked validation, investigated allocative simulation and resource optimisation in the ICU, while no study investigated automatic monitoring or prognostication of COVID-19 patients. Belciug et al.’s study on ICU resource optimisation employed queueing theory, a mathematical field of study, and Artificial Immune Systems, an evolutionary AI algorithm that is uncommonly utilised in medical research. Unfamiliarity and the absence of general adoption of these methods within the medical community may contribute to the paucity of studies exploring less common but potentially impactful AI applications. As highlighted in previous literature ^19,41^, robust interdisciplinary collaboration and communication will be crucial in stimulating broader applications of AI for COVID-19 in intensive care and emergency settings, as well as the in medical literature at large.

Assessment of AI predictive models also revealed significant deficiencies in model development, validation, and reporting. Unfortunately, these findings corroborate with earlier systematic reviews on predictive models for COVID-19 ^19^ and in intensive care settings ^21^. Studies developing AI models should adhere to the TRIPOD reporting guidelines ^22^, PROBAST ^23^, or, ideally, recent AI-specific guidelines. These include the guidelines for transparency, reproducibility, ethics, and effectiveness (TREE) ^42^, CONSORT-AI (Consolidated Standards of Reporting Trials-Artificial Intelligence) ^43^, and SPIRIT-AI (Standard Protocol Items: Recommendations for Interventional Trials-Artificial Intelligence) ^44^. While the above guidelines provide comprehensive explanations and elaborations, we emphasise hereinafter several common problematic areas within the reviewed studies and recommendations for future studies.

The most common source of bias was an inadequate sample size, which was found in all studies. A low sample size introduces the risk of over-fitting and model optimism. A benchmark for the development of logistic regression models is 20 EPV ^4,23,45^, while models using AI algorithms like random forest, support vector machines, and neural networks may require up to 200 EPV to account for model optimism ^46^; a minimum of 100 EPV is recommended for validation studies ^23^. Missing data also contributed significantly to bias; only two studies appropriately handled and reported on missing data. Ideally, the proportion of missing data for each variable should be reported ^22^ and multiple imputation should be used to avoid bias from inappropriate exclusion of participants with missing data (i.e. complete-case analysis) ^47-50^. However, if complete-case analysis is used, authors should provide a comparative analysis of model performance with and without excluded participants to facilitate the judgement of bias from exclusion. For prognostic studies, studies often failed to appropriately account for censored patients (e.g. neither discharged nor admitted to the ICU). Censored patients should be handled using a time-to-event analysis such as Cox regression; inappropriate exclusion of these patients may lead to a skewed dataset that includes fewer patients without the outcome, introducing bias into the model ^23^. For diagnostic studies, bias was often introduced by using the reverse transcription-polymerase chain reaction (RT-PCR) test as the ground truth or gold-standard for COVID-19 diagnosis, despite potentially poor sensitivity ^51^. We recommend repeat RT-PCR testing to minimise the likelihood of false-negative tests in both diagnostic model development and validation studies.

Several key areas for improvements in reporting were identified in our study, including treatments received by participants, blinding, and study size determination. In particular, no study reported on calibration, a crucial yet often unevaluated measure of model performance ^52^. We recommend assessing calibration using the calibration hierarchy described by Van Calster et al. ^53^ instead of the commonly used Hosmer-Lemeshow test ^54^. This avoids artificial stratification of patients into risk groups and other limitations associated with the Hosmer-Lemeshow test ^52^.

Studies should also endeavour to validate their data using stricter validation techniques. Studies with smaller sample sizes should utilise re-sampling techniques, such as bootstrapping or *k*-fold cross-validation. Studies with larger sample sizes should use a non-random split of data (e.g. by location or time) or perform external validation on independent data, for example, from a different study ^22,23,55^. Validation using the same data for model development is inappropriate as it only provides apparent model performance. Similarly, validation using a random split of data, such as a ‘train-test’ split, has lower power than re-sampling techniques ^22,56^ and should be avoided.

In addition to the limitations in quality and reporting of AI applications, the narrow scope of applications being investigated naturally leads to fewer AI applications eventually being suitable for clinical use. While AI has been practically applied for the identification of candidate drugs for drug repurposing ^57^ and contact tracing ^18^, its application and utility for COVID-19 in clinical settings have been insignificant to date. Several studies have employed AI techniques for the detection and classification of COVID-19 images ^20^, however, none have been validated as a clinical diagnostic adjunct in the ED. Factors that may contribute to this lack of clinical validation include the high risk of bias within existing models ^19,58^, limited applicability of radiographic images for discriminating between multiple differential diagnoses, and the high prevalence of asymptomatic radiographs in patients who present soon after the onset of symptoms ^59,60^. Notwithstanding the high risk of bias and poor reporting of the reviewed AI models, AI algorithms tend to produce uninterpretable “black box” predictive models, which may lead to decreased acceptability of both diagnostic and prognostic AI applications amongst clinicians and hospital administrators. Some studies ^32,36,37^ have attempted to overcome this by using AI techniques for feature selection and presenting the final model as a decision tree or scoring system with clearly defined input variables. However, such simplifications of AI models curtail performance and limit the utility of the final model.

The above barriers to the validation and integration of AI in clinical settings may preclude significant contribution of AI to combatting the COVID-19 pandemic in intensive care units and emergency departments in the near future. However, improvements in the development, validation, and reporting of AI applications will be critical in advancing the applicability and acceptance of these systems in clinical settings in later phases of the COVID-19 pandemic and in future global health crises. Encouragingly, leading journals such as the *Lancet* family of journals have committed to enforcing AI-specific guidelines such as CONSORT-AI and SPIRIT-AI for submissions with an AI intervention ^61^. However, concerted effort is needed from the entire research community, including journals, editors, and authors, to normalise the use of these guidelines and checklists. Such changes will encourage improved development, reporting, and eventual clinical uptake of future AI applications.

### Limitations

The results from our systematic review should be considered along with the following limitations. Firstly, our search excluded non-peer-reviewed articles which may neglect the most recent literature but ensures a baseline quality of included studies. Secondly, we may have missed some relevant articles despite using a comprehensive search strategy due to publication in journals not indexed in the searched databases and variations in terminology used to describe AI algorithms and intensive care and emergency settings. We may also have missed AI applications that were deployed without publication in scientific literature; in particular, given the intense media attention and the pressure to deploy solutions quickly, AI solutions developed by governments and industry are more likely to be published in mass media formats rather than scientific journals. Thirdly, assessment according to PROBAST and, to a lesser extent, TRIPOD reporting guidelines still rely on a degree of subjectivity, despite comprehensive explanations and elaborations. Hence, other reviewers may arrive at slightly differing results. Lastly, the unprecedented volume of research on COVID-19 has resulted in a rapidly evolving body of literature. Hence, our findings are merely descriptive of the current state of affairs, which may change with welcome improvements and additions to the medical literature.

## 5 Conclusions

Despite widespread interest in novel technologies for the COVID-19 pandemic, our systematic review of the literature reveals that current AI applications were limited in both the range of applications and clinical applicability. Several significant issues in the development, validation, and reporting of AI applications undermine safe and effective implementation of these systems within intensive care units or emergency departments. The integration of new AI-specific reporting guidelines like CONSORT-AI and SPIRIT-AI into research and publication processes will be a vital step in creating future AI applications that are clinically acceptable in the current pandemic, future pandemics, and within the wider medical field. We also emphasise the importance of closer interdisciplinary collaboration between AI experts and clinicians.

## Data Availability

The datasets used and/or analysed during the current study are available from the corresponding author on reasonable request.

## 7 Author Contributions

NL conceived the study. MLC and NL designed the study. MLC and NL screened and reviewed the articles and extracted paper information. MLC and NL planned the formal analyses, analysed the data, and drafted the manuscript. MLC, MEHO, FJS, ZZ, SLL, AFWH, and NL made substantial contributions to results interpretation and critical revision of the manuscript. All authors read and approved the final manuscript.

## 9 Acknowledgements

Not applicable.

